# Comparison of body composition measures assessed by bioelectrical impedance analysis versus dual-energy X-ray absorptiometry in UK Biobank

**DOI:** 10.1101/2023.11.01.23297916

**Authors:** Qi Feng, Jelena Bešević, Megan Conroy, Wemimo Omiyale, Ben Lacey, Naomi Allen

## Abstract

**Background:** Bioelectrical impedance analysis (BIA) and dual-energy X-ray absorptiometry (DXA) are used commonly for assessing body composition. This study aimed to evaluate the agreement between BIA and DXA measures to assess the validity of BIA measures in UK Biobank.

**Methods:** UK Biobank participants with body fat mass (FM) and fat free mass (FFM) estimates derived from BIA and DXA measures performed on the same date were included. BIA and DXA-derived estimates were compared using Pearson correlation coefficients. Bland-Altman analysis was performed to quantify the differences and its distribution. Multivariable linear regression was used to identify predictors for the difference between the two measures. Finally, prediction models were developed to calibrate BIA measures against DXA measures.

**Results:** 34437 participants (female 51.4%, mean age 55.2 years) were included. BIA and DXA measurements were highly correlated (correlation coefficient 0.96 for FM and 0.97 for FFM), with similar values in males and females and across body mass index subgroups. BIA underestimates FM by 1.84 kg (23.77 vs. 25.61), but overestimated FFM by 2.56 kg (52.49 vs. 49.93). The BIA-DXA difference was associated with individual FM, FFM, BMI and waist circumference. Prediction models showed overall good model performance in the training and testing data.

**Conclusion:** We found good agreement between BIA-and DXA-derived body composition measures at a population level in UK Biobank. However, BIA-DXA difference existed at individual level, and was associated with anthropometric measures. Future studies may consider using prediction models to calibrate BIA measures.

## Introduction

Body composition measures, which estimate the amount of body fat mass (FM) and fat free mass (FFM), are closely related to individual nutritional status and fitness, and have been recognised as risk factors for cardiovascular diseases and premature mortality ^1–3^. Commonly used automated techniques for measuring body composition include bioelectrical impedance analysis (BIA) and dual-energy X-ray absorptiometry (DXA). BIA measures the impedance of the water compartment of the human body to electrical currents, from which FM and FFM are predicted based on specific algorithms ^4^. During DXA assessment, the human body is scanned with X-rays of two different levels of energy and partitioned into FM, lean mass and bone mineral mass ^5^. DXA is usually used as the reference method for its high accuracy and precision ^6^. However, because BIA is easier to use, quicker, cheaper and does not involve exposure to radiation, it is more feasible to implement for large-scale studies.

Previous studies have examined the agreement between BIA-and DXA-derived body composition measures, but results are inconsistent by sex and across different BMI groups. Wan *et al.* observed a high correlation between BIA-and DXA-measured FFM, and found that BIA slightly overestimated FFM in overweight adolescents ^7^. Among patients followed up for malnutrition, obesity or eating disorders, BIA overestimated FFM and underestimated FM in people with body mass index (BMI) between 18.5 and 40, whereas the opposite was observed in people with BMI < 16 ^8^. Steiner *et al.* found that BIA underestimated FFM in males with chronic obstructive pulmonary disease while overestimated FFM in female counterparts ^9^. Leahy *et al.* found that BIA’s overestimation of FFM and underestimation of FM was similar in young males and females aged 18 to 30 years old ^10^.

UK Biobank is a prospective cohort of half million participants aged 40-69 years old at recruitment ^11^. Since 2014, the UK Biobank Imaging Study has been performing imaging scans in up to 100 000. During this assessment, participants undergo a BIA and DXA scan for body composition assessment. This provides a valuable opportunity to evaluate the accuracy of BIA-derived body composition measures against DXA in UK Biobank.

## Participants and methods

### Participants

UK Biobank is a cohort of half million participants aged 40-69 years old enrolled across England, Scotland and Wales during 2006-2010. Baseline measures included socioeconomic status, anthropometry, lifestyle, health status, medication and medical history, along with a range of physical measures and biological sampling. UK Biobank was approved by the North West Multicenter Research Ethics Committee. Since 2014, up to 100 000 participants are taking part in an Imaging Study. To date, DXA and BIA body composition data for about 50 000 participants have been made available.

### DXA

DXA was performed with Lunar iDXA Scanner (General Electric Healthcare, Wisconsin US). The scanner was calibrated daily following standard quality control procedures to maintain consistency. Participants were asked to lie flat on their back on the scanning couch for whole body scan. All operators had standardised central training, and all scans were performed according to standard operating procedures. Each scan acquisition was assessed in real time by the technicians for completeness, movement artefact and presence of any foreign bodies.

### BIA

BIA was performed with Tanita BC418MA Body Composition Analyser (Tanita, Japan). The analyser measured body impedance with a high frequency current (50 kHz) and 8 contact electrodes. Participants were asked to place their bare feet on the analyser platform, to keep their feet still and in good contact with the platform’s metallic electrodes on platform, and to grip the two handles firmly with palm and thumb contacting metallic electrodes and arms hanging loosely by their sides.

### Other measurements

Height was measured using a telescopic height rod device Seca 202 (Girodmedical, UK). Townsend Deprivation Index is a postcode-derived measure of socioeconomic deprivation status. Educational attainment was categorised into lower than university, university and above. Ethnicity included White, Black, Asian, Mixed and others. Smoking and drinking status were coded as never, previous or current users. Body mass index (BMI) was calculated as body weight divided by height square. All measures were taken at the imaging assessment.

### Statistical analysis

The primary body composition measures were whole body FM and FFM (in kg). In this study, we included participants with available body composition data derived from BIA and DXA that passed quality control. We excluded participants that had any missing values for BIA or DXA-derived body weight, FM or FFM; that had a value of zero for any of these variables; that had implausible values for body weight, FM or FFM (e.g., weight < 20 kg); where the difference between body weight and the sum of FM and FFM was larger than 0.2 kg in DXA or BIA assessment; and for which the difference between BIA-and DXA-measured body weight was larger than 5% (relative to BIA measurement).

For descriptive statistics, we used the mean and standard deviation (SD) for continuous variables, and frequency and percentages for categorical variables.

We calculated Pearson’s correlation coefficients for BIA-and DXA-derived body composition measures. Scatter plots of DXA-derived FM and FFM with BIA were generated to visualise their relationship. The difference between BIA-and DXA-derived FM and FFM were compared using paired sample t test. Bland-Altman analysis was performed by plotting the differences between BIA and DXA (BIA - DXA) against their average value ^12^, from which the mean difference and its 95% limits of agreement were obtained to show the bias and its distribution. We performed subgroup analysis by sex and BMI categories (<25, 25-30, and ≥ 30 kg/m^2^).

To investigate the potential risk factors for the difference between the BIA and DXA values, we performed multivariable linear regression and included age, BMI, height, waist circumference, DXA-derived FM and FFM in the model. We performed a sensitivity analysis by excluding participants with outlier measurements based on the criteria described in Malden et al.’s paper ^13^. Based on the identified risk factors, we developed linear models to calibrate BIA measures using DXA values (the “gold standard”), for each sex and BMI subgroups. In each subgroup, we randomly split the sample size into a training set and a testing set (80%:20%). R^2^ was estimated in the training set and the testing set to evaluate the overall goodness of fit. We fitted the predicted DXA values on the actual DXA measures in a linear model, to derive calibration slope and calibration-in-the-large (i.e., regression intercept) to evaluate the model calibration in the testing set. Ideally, calibration slope should be one and the calibration-in-the-large should be zero ^14^.

All analyses were performed in R software (version 4.1.1)

## Results

### Basic characteristics

Among the UK Biobank cohort, 34 437 participants (female 51.4%, age 55.2 [SD 7.6] years) were included in analysis (Supplementary figure 1). The baseline characteristics are shown in Table 1 and Supplementary table 1. The mean height and weight were 169.22 (9.28) cm and 76.25 (15.22) kg, respectively. The majority were White ethnicity (96.3%), 45.6% attended higher education, 95.2% were current drinkers and 6.4% current smokers. Males were more likely to be older, higher educated, and current drinkers and smokers.

**Table 1:**
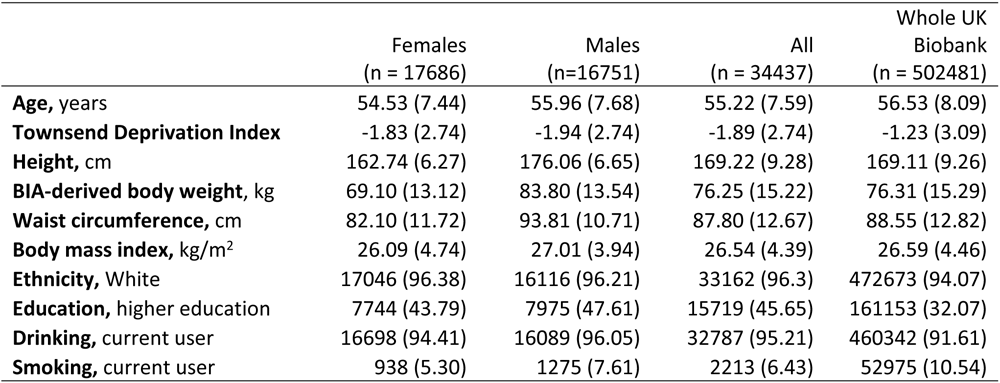
Basic characteristics of eligible participants and the whole UK Biobank cohort.

|  | Females<br>(n = 17686) | Males<br>(n=16751) | All<br>(n = 34437) | Whole UK<br>Biobank<br>(n = 502481) |
| --- | --- | --- | --- | --- |
| **Age, years** | 54.53 (7.44) | 55.96 (7.68) | 55.22 (7.59) | 56.53 (8.09) |
| **Townsend Deprivation Index** | -1.83 (2.74) | -1.94 (2.74) | -1.89 (2.74) | -1.23 (3.09) |
| **Height, cm** | 162.74 (6.27) | 176.06 (6.65) | 169.22 (9.28) | 169.11 (9.26) |
| **BIA-derived body weight, kg** | 69.10 (13.12) | 83.80 (13.54) | 76.25 (15.22) | 76.31 (15.29) |
| **Waist circumference, cm** | 82.10 (11.72) | 93.81 (10.71) | 87.80 (12.67) | 88.55 (12.82) |
| **Body mass index, kg/m<sup>2</sup>** | 26.09 (4.74) | 27.01 (3.94) | 26.54 (4.39) | 26.59 (4.46) |
| **Ethnicity, White** | 17046 (96.38) | 16116 (96.21) | 33162 (96.3) | 472673 (94.07) |
| **Education, higher education** | 7744 (43.79) | 7975 (47.61) | 15719 (45.65) | 161153 (32.07) |
| **Drinking, current user** | 16698 (94.41) | 16089 (96.05) | 32787 (95.21) | 460342 (91.61) |
| **Smoking, current user** | 938 (5.30) | 1275 (7.61) | 2213 (6.43) | 52975 (10.54) |

### Correlations between BIA and DXA

BIA and DXA body composition measures were highly correlated, with a correlation coefficient of 0.96 for FM and 0.97 for FFM (Table 2). The correlation between BIA and DXA for FFM was slightly higher in females than in males (0.98 vs. 0.92), but similar across BMI categories. By contrast, the correlation for FM was identical in males and females (0.97), but varied across BMI categories, with a higher correlation in participants with BMI < 25 in both sexes. Scatter plots similarly showed good agreement (R^2^ > 99% for all subgroups), but suggested that BIA overestimated FFM in males and females, but underestimated FM, especially in males (Figure 1). Sensitivity analysis excluding outliers showed similar results (Supplementary figure 2).

**Figure 1:**
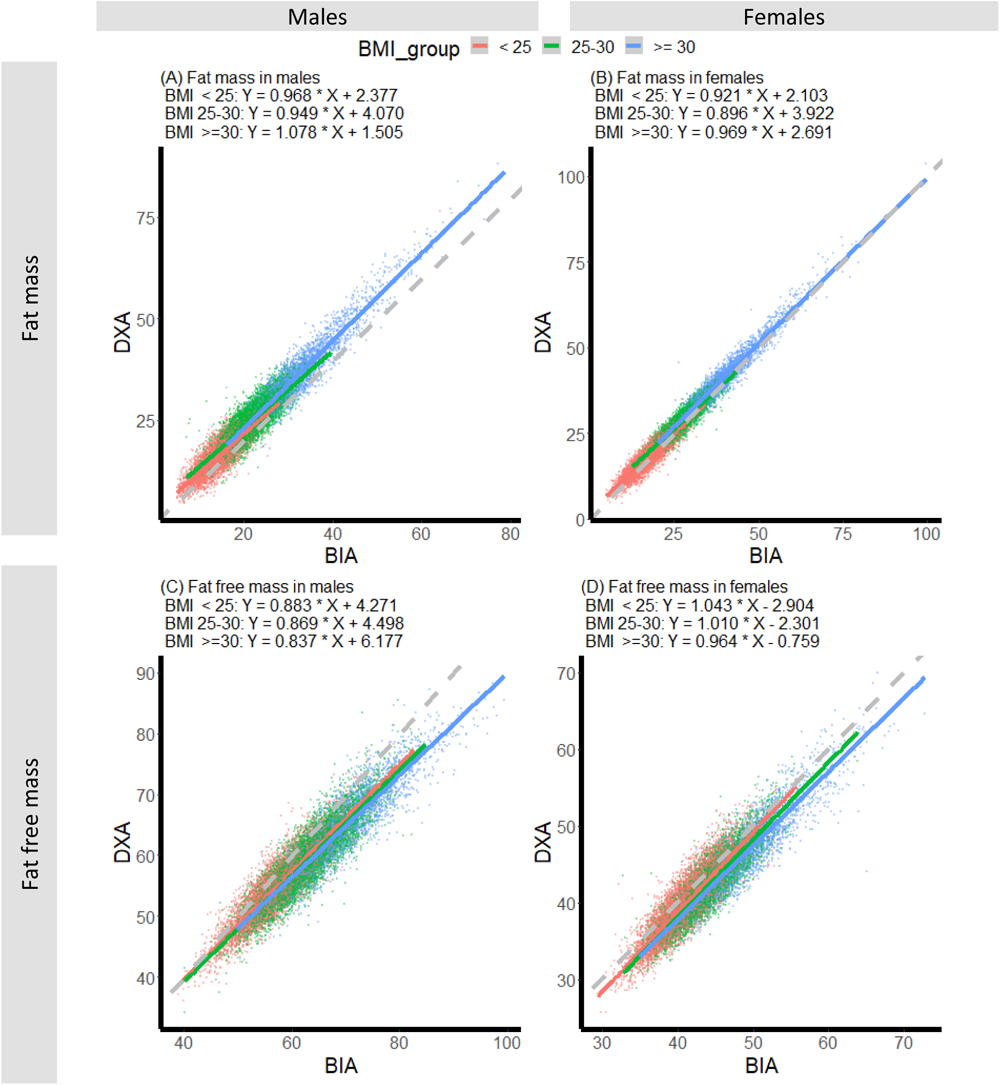
Scatter plots of BIA-and DXA-measured fat mass (kg) and fat free mass (kg). The colored solid lines are the linearly fitted lines. The gray dashed lines are reference lines (y = x), indicating perfect agreement between BIA and DXA. BIA: bioelectrical impedance analysis. DXA: dual-energy X-ray absorptiometry.

**Table 2:**
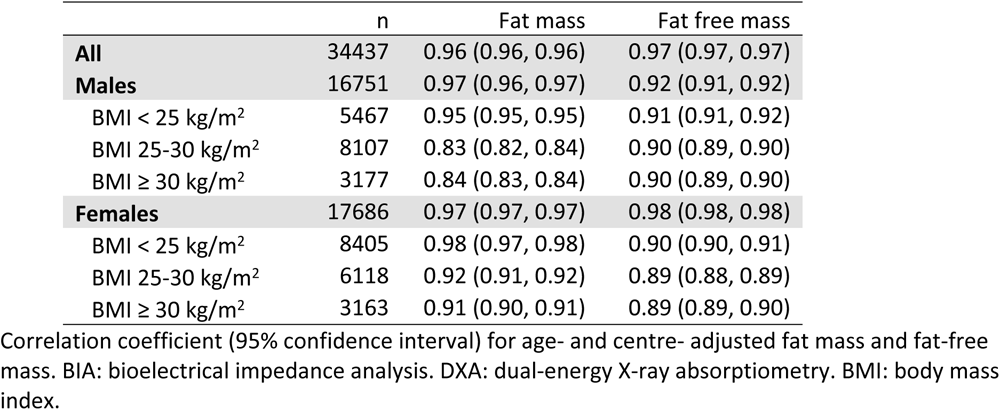
Pearson correlation coefficient between BIA-measured and DXA-measured fat mass and fat-free mass.

### Differences between BIA and DXA

The overall BIA-derived FM was lower than DXA (23.77 vs. 25.61, difference -1.84 kg), while BIA-derived FFM was higher than DXA (52.49 vs. 49.93, difference 2.56 kg), which was consistent in all sex and BMI subgroups. BIA-DXA differences were larger in males than females for both FM and FFM (-2.81 kg vs. -0.92 kg for FM, 3.54 kg vs. 1.64 kg for FFM, respectively). BIA underestimated FM by 11.39% in males and 3.47% in females, which was consistent across BMI subgroups. In contrast, BIA overestimated FFM by 6.06% and 3.92% in males and females, respectively, with smaller difference in participants with lower BMI (Table 3 and Figure 2).).

**Figure 2:**
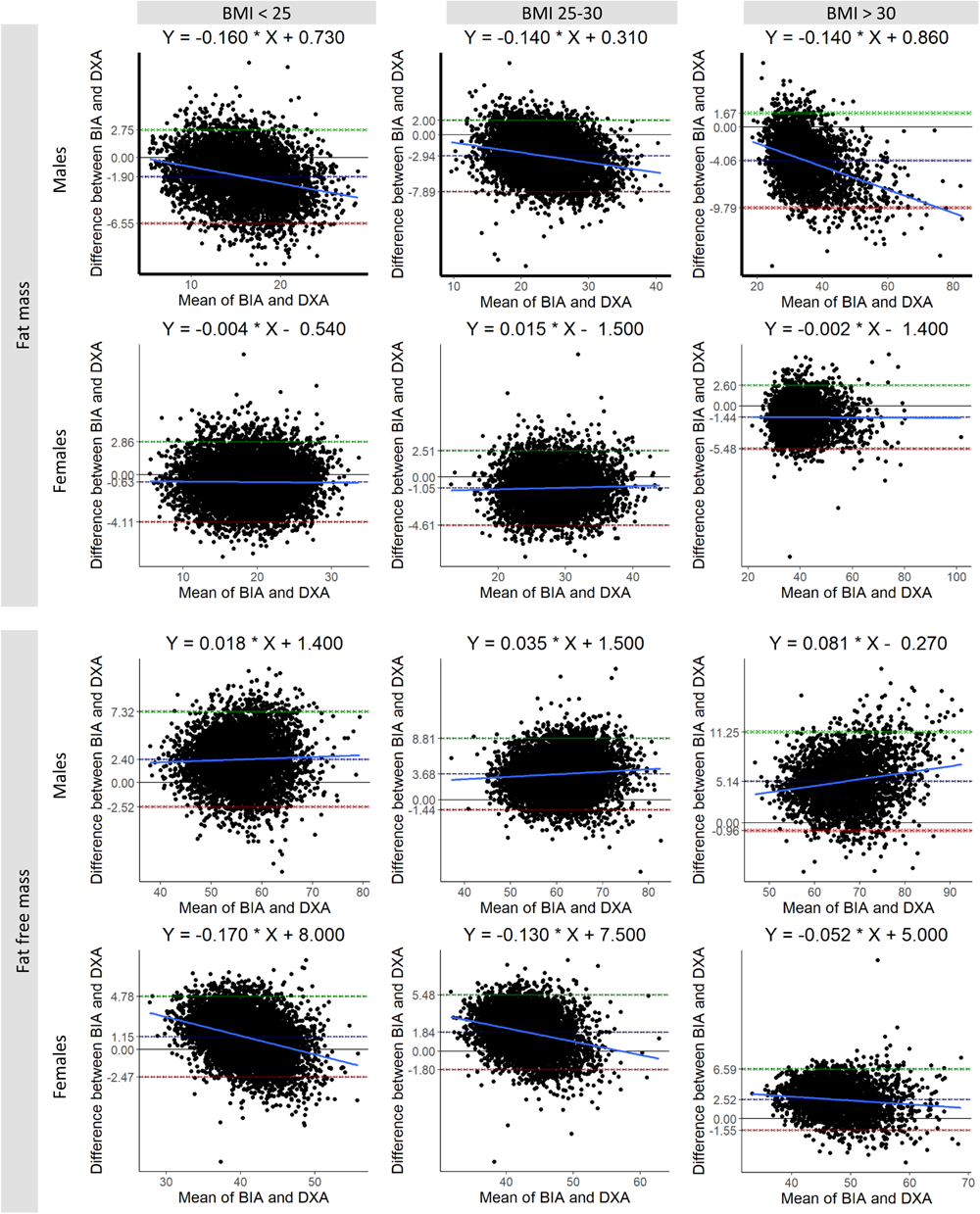
Bland-Altman plots for BIA-and DXA-measured fat mass (kg) and fat free mass (kg) in males and females. Showing the average bias and its 95% limits of agreement. Shaded band: 95% confidence interval. BIA: bioelectrical impedance analysis. DXA: dual-energy X-ray absorptiometry.

**Table 3:** Comparison of fat mass (kg) and fat free mass (kg) obtained by BIA versus by DXA.

|  | n | BIA | DXA | Difference (95%CI) | Difference % | p | 95% LOA |
| --- | --- | --- | --- | --- | --- | --- | --- |
| **Fat mass** |  |  |  |  |  |  |  |
| **All** | 34437 | 23.77 (8.81) | 25.61 (9.25) | -1.84 (-1.87, -1.81) | 7.18 | <0.01 | -6.70, 3.02 |
| **Males** | 16751 | 21.85 (7.80) | 24.67 (8.82) | -2.81 (-2.86, -2.77) | 11.39 | <0.01 | -8.04, 2.41 |
| BMI < 25 kg/m<sup>2</sup> | 5467 | 15.04 (3.68) | 16.95 (4.28) | -1.90 (-1.96, -1.84) | 11.21 | <0.01 | -6.55, 2.75 |
| BMI 25-30 kg/m<sup>2</sup> | 8107 | 22.11 (4.04) | 25.05 (4.58) | -2.94 (-3.00, -2.89) | 11.74 | <0.01 | -7.89, 2.00 |
| BMI ≥ 30 kg/m<sup>2</sup> | 3177 | 32.91 (7.32) | 36.97 (8.39) | -4.06 (-4.16, -3.96) | 10.98 | <0.01 | -9.79, 1.67 |
| **Females** | 17686 | 25.59 (9.31) | 26.50 (9.56) | -0.92 (-0.95, -0.89) | 3.47 | <0.01 | -4.58, 2.75 |
| BMI < 25 kg/m<sup>2</sup> | 8405 | 18.79 (4.36) | 19.41 (4.38) | -0.63 (-0.66, -0.59) | 3.25 | <0.01 | -4.11, 2.86 |
| BMI 25-30 kg/m<sup>2</sup> | 6118 | 27.55 (4.29) | 28.60 (4.23) | -1.05 (-1.10, -1.00) | 3.67 | <0.01 | -4.61, 2.51 |
| BMI ≥ 30 kg/m<sup>2</sup> | 3163 | 39.84 (8.04) | 41.28 (8.05) | -1.44 (-1.51, -1.37) | 3.49 | <0.01 | -5.48, 2.60 |
| **Fat free mass** |  |  |  |  |  |  |  |
| **All** | 34437 | 52.49 (11.13) | 49.93 (10.14) | 2.56 (2.53, 2.59) | 5.13 | <0.01 | -2.56, 7.69 |
| **Males** | 16751 | 61.96 (7.47) | 58.42 (6.75) | 3.54 (3.50, 3.58) | 6.06 | <0.01 | -2.05, 9.13 |
| BMI < 25 kg/m<sup>2</sup> | 5467 | 57.00 (5.62) | 54.61 (5.52) | 2.40 (2.33, 2.46) | 4.39 | <0.01 | -2.52, 7.32 |
| BMI 25-30 kg/m<sup>2</sup> | 8107 | 62.38 (5.89) | 58.70 (5.70) | 3.68 (3.63, 3.74) | 6.27 | <0.01 | -1.44, 8.81 |
| BMI ≥ 30 kg/m<sup>2</sup> | 3177 | 69.43 (7.28) | 64.29 (6.74) | 5.14 (5.04, 5.25) | 8.00 | <0.01 | -0.96, 11.25 |
| **Females** | 17686 | 43.52 (4.81) | 41.89 (4.92) | 1.64 (1.61, 1.66) | 3.92 | <0.01 | -2.21, 5.49 |
| BMI < 25 kg/m<sup>2</sup> | 8405 | 41.07 (3.43) | 39.92 (4.02) | 1.15 (1.11, 1.19) | 2.88 | <0.01 | -2.47, 4.78 |
| BMI 25-30 kg/m<sup>2</sup> | 6118 | 44.00 (3.64) | 42.15 (4.12) | 1.84 (1.80, 1.89) | 4.37 | <0.01 | -1.80, 5.48 |
| BMI ≥ 30 kg/m<sup>2</sup> | 3163 | 49.12 (4.95) | 46.60 (5.20) | 2.52 (2.45, 2.59) | 5.41 | <0.01 | -1.55, 6.59 |

Difference = BIA – DXA. P: p value for paired sample t test. Difference % = difference / DXA measurement. LOA: limits of agreement based on Bland-Altman analysis. BIA: bioelectrical impedance analysis. DXA: dual-energy X-ray absorptiometry. BMI: body mass index.

Bland-Altman plots showed similar patterns, but there was wide variation in the mean difference at individual level, demonstrated by the wide 95% limits of agreement. For FM, BIA-DXA difference was associated with individual FM in males across all BMI categories, but the association was much weaker in females. For FFM, the association between BIA-DXA difference and individual FFM was positive in males, but negative in females. However, the strength of the associations varied across BMI categories. (Figure 2)

Table 4 shows the results of multivariable regressions on BIA-DXA differences. These variables together explained 34.9-46.2% of the variation, with a higher proportion explained in males than in females. After adjusting for age, BMI, waist circumference and height, individual FM and FFM were still associated with BIA-DXA differences. More specifically, FM was negatively associated with the difference in FM in both male (beta [95% confidence interval]: -0.37 [-0.40, -0.34]) and females (-0.25 [-0.27, -0.23]), while FFM was positively associated (0.06 [0.03, 0.09] for males and 0.13 [0.11, 0.15] for females). For difference in FFM, FFM showed negative association in male (-0.37 [-0.39, -0.34]) and females (-0.36 [-0.38, -0.34]), but FM showed positive association (0.11 [0.08, 0.14] for males and 0.06 [0.04, 0.08] for females). BIA’s underestimation of FM was smaller in people with low FM, but the overestimation of FFM was smaller in people with high FFM. BMI showed consistent positive associations with differences in FM and in FFM, in males and in females. Waist circumference was positively associated with FM and FFM, consistent in both sexes.

**Table 4:** Results of multivariable linear regression of the BIA-DXA difference.

|  | BIA-DXA difference in fat mass |  |  | BIA-DXA difference in fat free mass |  |  |
| --- | --- | --- | --- | --- | --- | --- |
|  | Coefficient (95%CI) | P value | R<sup>2</sup>* | Coefficient (95%CI) | P value | R<sup>2</sup>* |
| **Male** |  |  |  |  |  |  |
| DXA-derived fat mass | -0.37 (-0.40, -0.34) | <0.01 | 39.4% | 0.11 (0.08, 0.14) | <0.01 | 46.2% |
| DXA-derived fat free mass | 0.06 (0.03, 0.09) | <0.01 |  | -0.37 (-0.39, -0.34) | <0.01 |  |
| BMI | 0.36 (0.27, 0.45) | <0.01 |  | 0.54 (0.45, 0.63) | 0.01 |  |
| Waist circumference | 0.05 (0.04, 0.05) | <0.01 |  | -0.05 (-0.05, -0.04) | <0.01 |  |
| Height | -0.01 (-0.03, 0.03) | 0.64 |  | 0.29 (0.26, 0.31) | <0.01 |  |
| **Female** |  |  |  |  |  |  |
| DXA-derived fat mass | -0.25 (-0.27, -0.23) | <0.01 | 34.8% | 0.06 (0.04, 0.08) | <0.01 | 40.72% |
| DXA-derived fat free mass | 0.13 (0.11, 0.15) | <0.01 |  | -0.36 (-0.38, -0.34) | <0.01 |  |
| BMI | 0.28 (0.22, 0.34) | <0.01 |  | 0.29 (0.23, 0.34) | <0.01 |  |
| Waist circumference | 0.03 (0.03, 0.04) | <0.01 |  | -0.03 (-0.03, -0.03) | <0.01 |  |
| Height | 0.09 (0.07, 0.11) | <0.01 |  | 0.09 (0.08, 0.11) | <0.01 |  |

All variables were fitted as continuous variables. Models were adjusted for age. BIA: bioelectrical impedance analysis. DXA: dual-energy X-ray absorptiometry. CI: confidence interval. BMI: body mass index. \*: R<sup>2</sup> of the fitted model.

Using DXA measures as gold standard, we developed linear models to calibrate BIA values for each sex and BMI subgroup (Table 5). Overall, the prediction models yielded high R^2^ for FM and FFM prediction, similar in training and testing sets, although variation existed. FM prediction models showed higher R^2^ for females than for males, and the highest R^2^ was seen in the the subgroup with BMI > 30. The variation of R^2^ across BMI subgroups seemed smaller for FFM prediction models than for FM models. Most models demonstrated good calibration in the testing sets, as we observed no evidence for calibration slopes deviating from one and for calibration-in-the-large deviating from zero.

**Table 5:** Prediction equations for DXA-measured fat mass and fat-free mass by sex and BMI.

|  |  |  | Training | Testing |  |  |
| --- | --- | --- | --- | --- | --- | --- |
| Sex | BMI | Prediction equation | R<sup>2</sup> (%) | R<sup>2</sup> (%) | Calibration slope | Calibration-in-the-large |
|  |  | FM_DXA |  |  |  |  |
| Male | < 25 | FM_DXA = -2.715 + 0.869 * FM_BIA + 0.150 * FFM_BIA + 0.105 * WC + -0.061 * height | 73.13 | 74.24 | 1.01 (0.97, 1.04) | -0.15 (-0.77, 0.46) |
|  | 25 - 30 | FM_DXA = -5.046 + 0.832 * FM_BIA + 0.162 * FFM_BIA + 0.123 * WC + -0.057 * height | 74.86 | 73.81 | 0.98 (0.95, 1.01) | 0.50 (-0.21, 1.22) |
|  | > 30 | FM_DXA = -5.908 + 0.909 * FM_BIA + 0.163 * FFM_BIA + 0.115 * WC + -0.061 * height | 89.88 | 90.86 | 0.96 (0.94, 0.99) | 1.13 (0.23, 2.04) |
| Female | < 25 | FM_DXA = 16.291 + 0.954 * FM_BIA + 0.119 * FFM_BIA + 0.011 * WC + -0.126 * height | 85.78 | 85.61 | 1.00 (0.99, 1.02) | 0.00 (-0.39, 0.39) |
|  | 25 - 30 | FM_DXA = 16.446 + 0.957 * FM_BIA + 0.186 * FFM_BIA + 0.012 * WC + -0.145 * height | 83.93 | 85.09 | 1.01 (0.99, 1.03) | -0.25 (-0.93, 0.43) |
|  | > 30 | FM_DXA = 18.079 + 0.952 * FM_BIA + 0.213 * FFM_BIA + -0.023 * WC + -0.141 * height | 94.32 | 94.55 | 0.99 (0.97, 1.01) | 0.34 (-0.44, 1.12) |
|  |  | FFM_DXA |  |  |  |  |
| Males | < 25 | FFM_DXA = 3.807 + 0.114 * FM_BIA + 0.857 * FFM_BIA + -0.120 * WC + 0.059 * height | 81.61 | 83.23 | 1.03 (1.00, 1.05) | -1.38 (-2.88, 0.12) |
|  | 25 - 30 | FFM_DXA = 6.152 + 0.141 * FM_BIA + 0.842 * FFM_BIA + -0.129 * WC + 0.052 * height | 82.07 | 82.26 | 1.01 (0.99, 1.03) | -0.64 (-1.98, 0.70) |
|  | > 30 | FFM_DXA = 7.017 + 0.051 * FM_BIA + 0.839 * FFM_BIA + -0.118 * WC + 0.057 * height | 82.86 | 83.53 | 1.02 (0.99, 1.06) | -1.15 (-3.40, 1.11) |
| Female | < 25 | FFM_DXA = -16.697 + 0.021 * FM_BIA + 0.880 * FFM_BIA + -0.015 * WC + 0.130 * height | 81.42 | 80.46 | 0.98 (0.96, 1.01) | 0.50 (-0.42, 1.43) |
|  | 25 - 30 | FFM_DXA = -17.133 + 0.020 * FM_BIA + 0.814 * FFM_BIA + -0.012 * WC + 0.148 * height | 81.72 | 83.37 | 1.00(0.98, 1.03) | -0.08 (-1.13, 0.98) |
|  | > 30 | FFM_DXA = -18.250 + 0.023 * FM_BIA + 0.798 * FFM_BIA + 0.020 * WC + 0.141 * height | 85.58 | 87.49 | 1.03 (1.00, 1.06) | -1.30 (-2.71, 0.13) |

BMI: body mass index. FM: fat mass. FFM: fat free mass. DXA: BIA: bioimpedance analysis. WC: waist circumference. RMSE: root mean squared error. In each subgroup, individuals were randomly assigned to a training set and a testing set (80%:20%). Prediction equations were derived from the training sets, and validated in the testing sets. Calibration slope and calibration-in-the-large: observed values were fitted on predicted values in a linear regression model, from which the slope is the calibration slope, and the intercept is the calibration-in-the-large. Ideally, calibration slope should be 1 and calibration-in-the-large should be 0.

## Discussion

The present study demonstrated overall high correlation between BIA and DXA in measuring body FM and FFM in UK Biobank, although small differences existed at an individual level. Compared with DXA, BIA underestimated FM by an average of 1.84 kg, and it overestimated FFM by 2.56 kg. The difference between BIA and DXA was larger in males than in females, and was additionally associated with body composition and other anthropometric measures.

The high correlation between BIA and DXA is consistent with some previous studies. For example, Pietilan *et al* ^15^ found a high correlation for FM between BIA and DXA over a one-year weight loss intervention program. We found that BIA underestimated FM but overestimated FFM, and the same pattern was consistently observed among all BMI subgroups. The overall difference in FM and FFM in our study was smaller than in Achamrah *et al*, which was more similar to the estimates in Sun *et al* ^16^ and Leahy *et al* ^10^.

We found that sex-specific BIA-DXA difference was associated with multiple factors, including BMI, waist circumference, as well as body composition measures. These factors explained more variance in BIA-DXA difference in males than in females. After adjustment, BIA’s underestimation of FM was smaller in people with low FM, but the overestimation of FFM was smaller in people with high FFM. Sun et al ^16^ found that BIA-DXA difference in FM percentage was negatively associated with DXA-derived FM percentage. Our analysis also showed that BMI and waist circumference was negatively associated with the FM difference between BIA and DXA. Therefore, the FM difference would be reduced in males who have high BMI, high waist circumference and low FM, and in the females who additionally have higher FFM and height.

BIA analyser is based on the principle that electric current flows at different rates through the body, depending upon its composition (FM and FFM). It measures body impedance, and predicts FM percentage based on regression equations, and finally translates FM percentage to FM and FFM in kg ^17^. Therefore, the selection of equation is important. The exact in-built predictive equation used by the BIA analyser in UK Biobank (Tanita BC418) is not clear, but the equation was derived from a case-mix of Japanese and western populations (according to the device manual). It has been shown that BIA performs better in a population of single ethnicity, than in a complex multi-ethnic population ^17^. The ethnic composition of these UK biobank analyses are British White (about 95%), which increases the reliability of body composition estimates derived by the BIA device. The same device mode (Tanita BC418) has been used and compared against DXA in previous studies, which yielded inconsistent results ^18–21^. This might be due to the differences in participants’ age, sex, body size and body composition, which we have shown to influence the results.

The high correlation and small average difference between the two methods indicate that BIA is a good surrogate of DXA at population level in UKB. In future epidemiological studies of body composition, researchers could consider using either BIA-measured FM and FFM or calibrated values based on BIA measures and the prediction models developed in this study, which would substantially increase sample size and statistical power while maintaining high accuracy of measurement. Compared with DXA, BIA has logistic advantages of low cost, rapidity, high accessibility, convenience and free of radiation. When using BIA for body composition assessment, multiple other factors may influence the results, including changes in quantity and distribution of body water (diet, exercise, oedema of distal extremities, etc.), body skin temperature, ambient temperature, contacts between limbs and trunk ^17^.

However, differences between the two methods exist at individual level, as suggested by the wide limits of agreements. Bland-Altman analysis showed that 95% subjects would have BIA-DXA difference of FM between -8.04 to 2.41 in males and -4.58 to 2.75 in females. This indicates that for personal health monitoring and clinical practice, using BIA for body composition may cause bias to a certain degree. In this case, the prediction models that calibrate BIA measures against DXA measures should be useful. Although BIA is not an absolutely accurate alternative for DXA for cross-sectional comparison, studies have shown that BIA can assess FM changes longitudinally as well as DXA ^15^.

This study has some limitations. First, the analysis was based on data from about 35000 participants in UK Biobank, because the recruitment in the imaging study is still ongoing. The current sample size has sufficiently powered to detect a correlation coefficient > 0.8 (power > 99.9%). Compared with the whole UK Biobank cohort, our sample is slightly younger, but the body size and body composition measures are comparable. Second, we did not compare lean mass. Because BIA is a two-compartment model, in which human body is partitioned into FM and FFM. By contrast, DXA is a three-compartment model, in which FFM is further partitioned into lean mass and bone mineral mass. Therefore, BIA FFM is not comparable to DXA lean mass. Third, we did not compare segmental body composition, such as trunk, limbs and visceral parts. In UK Biobank, BIA provided segmental FFM for different body regions, while DXA only have lean mass for different body regions. Fourth, we did not evaluate the agreement between BIA and DXA in monitoring changes in body composition longitudinally in this study, due to the cross-sectional design.

## Conclusions

This study of 35000 UK Biobank participants revealed high correlation and good agreement between BIA-and DXA-derived body composition measures at population level. Overall, BIA is a good surrogate for DXA; future studies of UK Biobank can use BIA-derived body composition, to increase sample size. BIA-DXA difference was associated with age, sex, and body composition.

## Ethics approval

The Research Tissue Bank approval from its governing Research Ethics Committee was obtained for the UK Biobank, as recommended by the National Research Ethics Service. Permission to use the UK Biobank Resource was approved by the access subcommittee of the UK Biobank Board. Written informed consent was obtained for all participants.

## Data sharing statement

The data that support the findings of this study are available from the UK Biobank but restrictions apply to the availability of these data, which were used under license for the current study, and so are not publicly available. The UK Biobank resources are however available and can be accessed through applications on their website (www.ukbiobank.ac.uk/). Analytic codes are available upon reasonable request.

## Conflict of interest

There is no conflict of interest.

## Sources of financial support

QF, JB, MC, WO, BL and NA are jointly supported by Wellcome Trust, UKRI, BHF, CRUK and NIHR [223600/Z/21/Z]. The funder of the study had no role in study design, data collection, data analysis, data interpretation, writing of the report or involved in the decision to submit the paper for publication.

## Data Availability

The data that support the findings of this study are available from the UK Biobank but restrictions apply to the availability of these data, which were used under license for the current study, and so are not publicly available. The UK Biobank resources are however available and can be accessed through applications on their website (https://www.ukbiobank.ac.uk/). Analytic codes are available upon reasonable request.

## Acknowledgement

The current research has been conducted using the UK Biobank Resource under the application No. 41115. We sincerely thank all UK Biobank participants and staff.

## Notes

### Competing Interest Statement

The authors have declared no competing interest.

### Funding Statement

The author(s) received no specific funding for this work.

